# Development of a Clinical Decision Support System for Severity Risk Prediction and Triage of COVID-19 Patients at Hospital Admission: an International Multicenter Study

**DOI:** 10.1101/2020.05.01.20053413

**Authors:** Guangyao Wu, Pei Yang, Henry C. Woodruff, Xiangang Rao, Julien Guiot, Anne-Noelle Frix, Michel Moutschen, Renaud Louis, Jiawei Li, Jing Li, Chenggong Yan, Dan Du, Shengchao Zhao, Yi Ding, Bin Liu, Wenwu Sun, Fabrizio Albarello, Alessandra D’Abramo, Vincenzo Schininà, Emanuele Nicastri, Mariaelena Occhipinti, Giovanni Barisione, Emanuela Barisione, Iva Halilaj, Yuanliang Xie, Xiang Wang, Pierre Lovinfosse, Jianlin Wu, Philippe Lambin

## Abstract

**Question:** How do nomograms and machine-learning algorithms of severity risk prediction and triage of COVID-19 patients at hospital admission perform?

**Findings:** This model was prospectively validated on six test datasets comprising of 426 patients and yielded AUCs ranging from 0.816 to 0.976, accuracies ranging from 70.8% to 93.8%, sensitivities ranging from 83.7% to 100%, and specificities ranging from 41.0% to 95.7%. The cut-off probability values for low, medium, and high-risk groups were 0.072 and 0.244.

**Meaning:** The findings of this study suggest that our models performs well for the diagnosis and prediction of progression to severe or critical illness of COVID-19 patients and could be used for triage of COVID-19 patients at hospital admission.

**IMPORTANCE:** The outbreak of the coronavirus disease 2019 (COVID-19) has globally strained medical resources and caused significant mortality for severely and critically ill patients. However, the availability of validated nomograms and the machine-learning model to predict severity risk and triage of affected patients is limited.

**OBJECTIVE:** To develop and validate nomograms and machine-learning models for severity risk assessment and triage for COVID-19 patients at hospital admission.

**DESIGN, SETTING, AND PARTICIPANTS:** A retrospective cohort of 299 consecutively hospitalized COVID-19 patients at The Central Hospital of Wuhan, China, from December 23, 2019, to February 13, 2020, was used to train and validate the models. Six cohorts with 426 patients from eight centers in China, Italy, and Belgium, from February 20, 2020, to March 21, 2020, were used to prospectively validate the models.

**MAIN OUTCOME AND MEASURES:** The main outcome was the onset of severe or critical illness during hospitalization. Model performances were quantified using the area under the receiver operating characteristic curve (AUC), accuracy, sensitivity, and specificity.

**RESULTS:** Of the 299 hospitalized COVID-19 patients in the retrospective cohort, the median age was 50 years ((interquartile range, 35.5-63.0; range, 20–94 years) and 137 (45.8%) were men. Of the 426 hospitalized COVID-19 patients in the prospective cohorts, the median age was 62.0 years ((interquartile range, 50.0-72.0; range, 19-94 years) and 236 (55.4%) were men. The model was prospectively validated on six cohorts yielding AUCs ranging from 0.816 to 0.976, with accuracies ranging from 70.8% to 93.8%, sensitivities ranging from 83.7% to 100%, and specificities ranging from 41.0% to 95.7%. The cut-off values of the low, medium, and high-risk probabilities were 0.072 and 0.244. The developed online calculators can be found at https://covid19risk.ai/.

**CONCLUSION AND RELEVANCE:** The machine learning models, nomograms, and online calculators might be useful for the prediction of onset of severe and critical illness among COVID-19 patients and triage at hospital admission. Further prospective research and clinical feedback are necessary to evaluate the clinical usefulness of this model and to determine whether these models can help optimize medical resources and reduce mortality rates compared with current clinical practices.

## Introduction

In December 2019, a novel coronavirus, severe acute respiratory syndrome coronavirus 2 (SARS-CoV-2; earlier named as 2019-nCoV), emerged in Wuhan, China.^1^ The disease caused by SARS-CoV-2 was named coronavirus disease 2019 (COVID-19). As of March 31, 2020, more than 750 000 COVID-19 patients have been reported globally, and over 36 000 patients have died.^2^ The outbreak of COVID-19 has developed into a pandemic.^3^

Among COVID-19 patients, around 80% present with mild illness whose symptoms usually disappear within two weeks.^4^ However, around 20% of the patients may proceed and necessitate hospitalization and increased medical support. The mortality rate for the severe patients is around 13.4%.^4^ Therefore, risk assessment of patients preferably in a quantitative, non-subjective way, is extremely important for patient management and medical resource allocation. General quarantine and symptomatic treatment at home or mobile hospital can be used for most non-severe patients, while a higher level of care and fast track to the intensive care unit (ICU) is needed for severe patients. Previous studies have summarized the clinical and radiological characteristics of severe COVID-19 patients,^5,6^ while the prognostic value of different variables is still unclear.

Machine-learning is a branch of artificial intelligence that learns from past data in order to build a prognostic model.^7^ In recent years, machine learning has been developed as a useful tool to analyze large amounts of data from medical records or images.^8^ Previous modeling studies focused on forecasting the potential international spread of COVID-19.^9^ However, to our knowledge, very few studies have used this approach to predict clinical outcomes of COVID-19 patients.

Therefore, our objective is to develop and validate a prognostic machine-learning model based on clinical, laboratory, and radiological variables of COVID-19 patients at hospital admission for risk assessment during hospitalization. Our ambition is to develop a multifactorial Decision Support Systems with different datasets to facilitate risk prediction and triage (home or mobile hospital quarantine, hospitalization, or ICU) of the patient at hospital admission.

## Methods

### Patients

The institutional review boards of The Central Hospital of Wuhan (No.2020-71) approved this study, which followed the Standards for Reporting of Diagnostic Accuracy Studies statement,^10^ and the requirement for written informed consent was waived. 299 adult confirmed COVID-19 patients from the central hospital of Wuhan were included consecutively and retrospectively between December 23, 2019 and February 13, 2020. The inclusion criteria were: (1) patients with a confirmed COVID-19 disease, (2) patients present at hospital for admission. The exclusion criteria were: (1) patients already with a severe illness at hospital admission; (2) time interval > 2 days between admission and examinations; and (3) no data available or delayed results as described below. The patients included from this center were divided into two datasets according to the entrance time of hospitalization, 80% for training (239 patients from December 23, 2019, to January 28, 2020) and 20% for internal validation (60 patients from January 29 to February 13, 2020). The test datasets were prospectively collected between February 20, 2020 and March 31, 2020 from other eight centers (**Supplement**) in China, Italy, and Belgium under the same inclusion and exclusion criteria (**Figure 1**).

**Figure 1.**
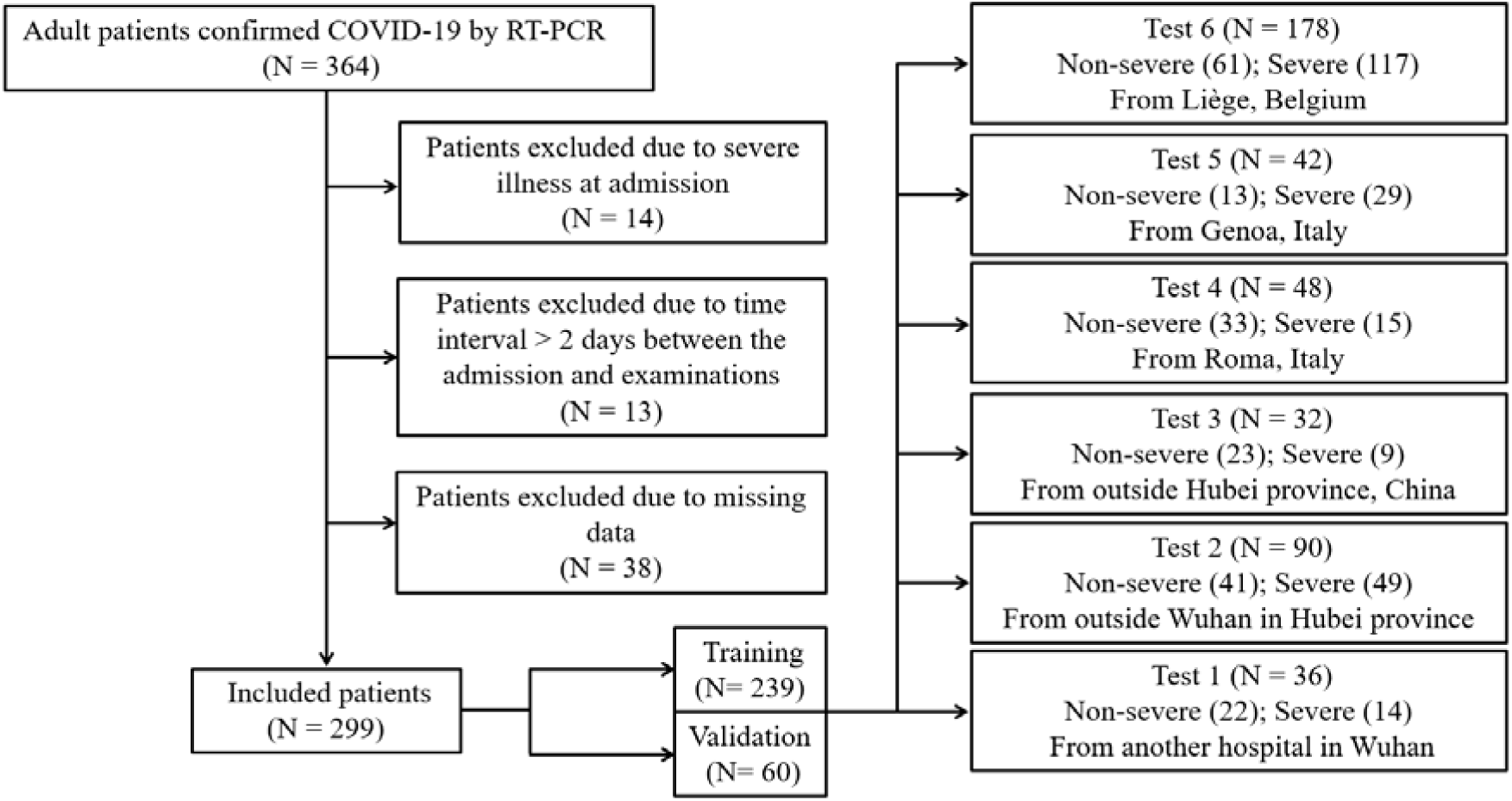
Flowchart of the patient selection process

Patients were labelled as having a “severe disease” if at least one of the following criteria were met during hospitalization:^11^ (1) respiratory distress with respiratory frequency ≥ 30/min; (2) pulse oximeter oxygen saturation ≤ 93% at rest; (3) oxygenation index (artery partial pressure of oxygen/inspired oxygen fraction) ≤ 300 mmHg; or (4) one of the following conditions occurs: (a) respiratory failure requiring mechanical ventilation; (b) shock; (c) ICU admission due to combined organ failure; or (d) death. Patients were labelled as having a “non-severe disease” if none of the above-mentioned criteria were met during the whole hospitalization process until deemed recovered and discharged from the hospital.

### Data collection

Clinical, laboratory, radiological characteristics and outcome data were obtained in the case record form shared by the International Severe Acute Respiratory and Emerging Infection Consortium from the electronic medical records.^12^ The clinical characteristics included basic information (5 variables), comorbidities (11 variables), and symptoms (13 variables). All clinical characteristics were obtained from the electronic medical system when the patients were admitted for the first time. A confirmed case with COVID-19 was defined as a positive result of high-throughput sequencing or real-time reverse-transcriptase polymerase-chain-reaction assay for nasal and pharyngeal swab specimens. 42 laboratory results were recorded, including complete blood count, white blood cell differential count, D-dimer, C-reactive protein (CRP), cardiac enzymes, procalcitonin, liver function test, kidney function test, B-type natriuretic peptide and electrolyte test. The arterial blood gas was not taken into account due to missing data for most early-stage patients. The metric conversion of laboratory results was performed using an online conversion table.^13^

The semantic CT characteristics (including ground-glass opacity, consolidation, vascular enlargement, air bronchogram, and lesion range score) were independently evaluated on all datasets by two radiologists (PY [a radiologist with 5 years’ experience in chest CT images] and YX [a radiologist with 20 years’ experience in chest CT images]), who were blinded to clinical and laboratory results. Any disagreement was resolved by a consensus read. Lesion range was identified as areas of ground-glass opacity or consolidation and was graded with a 6-point scale according to the lesion volume proportion in each single lobe: 0 = no lung parenchyma involved, 1 = up to 5% of lung parenchyma involved, 2 = 5-25%, 3 = 26-50%, 4 = 51-75%, and 5 = 76-100% of lung parenchyma involved. The lesion volume proportion was automatically calculated by Shukun Technology Pneumonia Assisted Diagnosis System (Version 1.17.0), and the final score is a total score from five lobes (**Figure 2**). Detailed CT acquisition and reconstruction parameters are presented in the **Supplement**.

**Figure 2.**
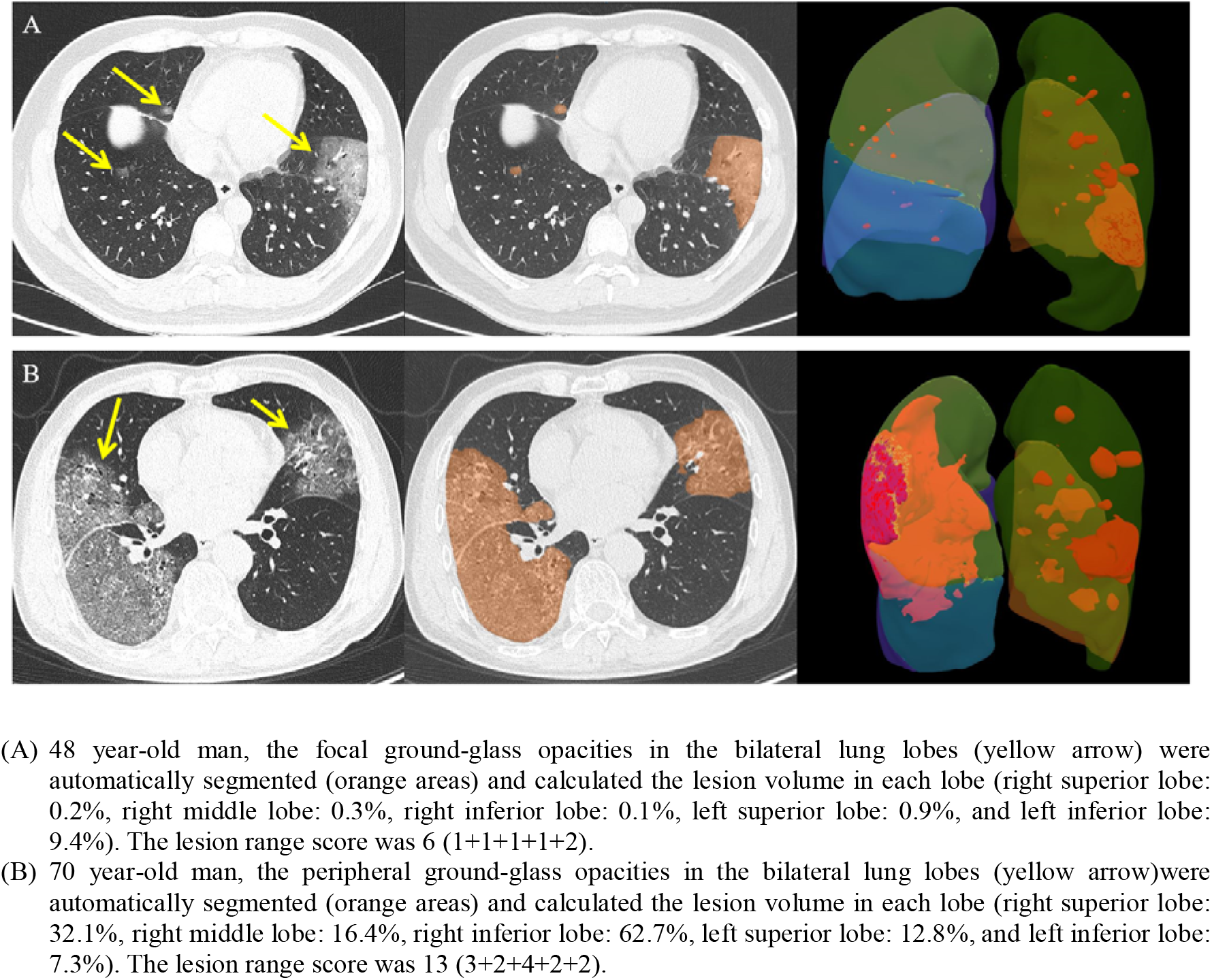
Chest CT images of two patients with COVID-19 pneumonia

### Feature selection and modeling

All feature selection and model training were performed in the training dataset alone to prevent information leakage. In order to reduce feature dimensionality, features showing high pairwise Spearman correlation (r > 0.8) and the highest mean correlation with all remaining features were removed, followed by application of the Boruta algorithm to select features with high importance and robustness.^14^ Recursive feature elimination based on bagged tree models with a cross-validation technique (10 folds, 10 times) was performed to select the best combination of features. The feature selection process was used for clinical, laboratory, and CT semantic models alone, and in combination.

Logistic regression models based on selected features were trained and the validation dataset was used to internally validate the prognostic performance of the models. Four models were trained: Model 1 contained only baseline clinical features before symptoms; Model 2 used all selected clinical features of symptomatic patients; Model 3 used selected semantic CT features and age; Model 4 employed all selected clinical, laboratory and CT features. A weight-balanced method was used during feature selection.

The prognostic performances of the best model were compared with other models on the training dataset, due to a bigger sample size. The performance of the best model was gauged on the six test datasets via the receiver operator characteristic (ROC), confusion matrix and calibration plot. In order to gauge the level of overfitting, the outcomes were randomized on the best model. The patients from the training and validation datasets were divided into low, medium and high risk according to the median +/- 25% interquartile range (IQR) of probabilities of the best model. The nomograms and on-line calculators were used to provide the interpretability of the trained models. The test datasets were used to gauge the prognostic performance and the validity of the risk cut-off values for the best model.

### Statistical analysis

Baseline data were summarized as median, and categorical variables as frequency (%). Differences between the severe group and the non-severe group were tested using the Mann-Whitney test for continuous data and Fisher’s exact test for categorical data. Feature correlations were measured using the Spearman correlation coefficient. We determined the area under the ROC curve (AUC) with its 95% confidence interval (CI) and tested AUC difference between Models 1-3 and Model 4 by the DeLong method,^15^ measures of prognostic performance included the AUC, accuracy, sensitivity and specificity. The accuracy value with 95% CI was obtained from the confusion-matrix. The calibration-plot was used to estimate the goodness-of-fit and consistency of the model on the test datasets. All p values were two-sided, and p < 0.05 was regarded as significant. All statistical analyses, modeling, and plotting were performed in R (version 3.5.3).

## Results

Of 299 hospitalized COVID-19 patients in retrospective cohort, the median age was 50 years ((interquartile range, 35.5-63.0; range, 20-94 years) and 137 (45.8%) were men. All the clinical characteristics and CT findings were summarized in **Table 1**, and more details of laboratory findings can be seen in **Table 2**. Of 426 hospitalized COVID-19 patients in prospective cohorts as test datasets, the median age was 62.0 years ((interquartile range, 50.0-72.0; range, 19-94 years) and 236 (55.4%) were men.

**Table 1.** Clinical characteristics and radiological findings of patients confirmed with COVID-19

|  | Non-severe group (n=217) | Severe group (n=82) | p value* |
| --- | --- | --- | --- |
| **Basic information** |  |  |  |
| Age | 42.0 (33.0-59.0) | 62.0 (53.0-71.8) | < .001 |
| No. of men | 90 (41.5) | 47 (57.3) | .019 |
| No. of smoker | 13 (6.0) | 5 (6.1) | 1 |
| No. of hospital staff | 86 (39.6) | 4 (4.9) | < .001 |
| Time of onset of illness, days | 4.0 (2.0- 7.0) | 4.0 (2.0-7.0) | .760 |
| **Comorbidities** |  |  |  |
| No. with hypertension | 36 (16.6) | 39 (47.6) | < .001 |
| No. with diabetes | 18 (8.3) | 19 (23.2) | .001 |
| No. with hyperlipidemia | 10 (4.6) | 6 (7.3) | .390 |
| No. with cardiopathy disease | 2 (0.9) | 8 (9.8) | < .001 |
| No. with chronic obstructive pulmonary disease | 7 (3.2) | 14 (17.1) | < .001 |
| No. with cerebrovascular disease | 6 (2.8) | 16 (19.5) | < .001 |
| No. with kidney disease | 4 (1.8) | 12 (14.6) | < .001 |
| No. with fatty liver | 25 (11.5) | 15 (18.3) | .131 |
| No. of Hepatitis B virus carrier | 2 (0.9) | 5 (6.1) | .018 |
| No. with cancer history | 13 (6.0) | 4 (4.9) | 1 |
| No. with surgical history | 26 (12.0) | 19 (23.2) | .019 |
| **Symptoms** |  |  |  |
| Fever | 162 (74.7) | 57 (69.5) | .382 |
| Body temperature, °C | 38.0 (37.1-38.5) | 37.8 (36.8-38.5) | .784 |
| Cough | 143 (65.9) | 56 (68.3) | .683 |
| Sputum | 56 (25.8) | 28 (34.1) | .194 |
| Weakness | 96 (44.2) | 37 (45.1) | 1 |
| Diarrhea | 24 (11.1) | 10 (12.2) | .839 |
| Vomiting | 15 (6.9) | 11 (13.4) | .105 |
| Chest tightness | 45 (20.7) | 35 (42.7) | < .001 |
| Dyspnoea | 9 (4.1) | 7 (8.5) | .152 |
| Muscular soreness | 59 (27.2) | 20 (24.4) | .662 |
| Chill | 35 (16.1) | 15 (18.3) | .729 |
| Conjunctival congestion | 1 (0.5) | 2 (2.4) | .184 |
| Headache or dizziness | 32 (14.8) | 14 (17.1) | .595 |
| **Radiological findings** |  |  |  |
| Main findings |  |  | .273 |
| Normal | 2 (0.9) | 3 (3.7) |  |
| Ground-glass opacity only | 127 (58.5) | 44 (53.7) |  |
| Consolidation only | 22 (10.1) | 6 (7.3) |  |
| Mixed | 66 (30.4) | 29 (35.4) |  |
| Vascular enlargement | 63 (29.0) | 39 (47.6) | .004 |
| Air-bronchogram | 45 (20.7) | 34 (41.5) | < .001 |
| Lesion range score | 4.0 (2.0-7.0) | 6.0 (4.0-10.8) | < .001 |

Data are median (IQR) and N (%) where N is the total number of patients with available data. p values comparing non-severe and severe groups were obtained Fisher's exact test or Mann-Whitney U test.

**Table 2.** Laboratory results of patients with COVID-19 at hospital admission

|  | Non-severe group (n=217) | Severe group (n=82) | p value* |
| --- | --- | --- | --- |
| **Complete blood cell count** |  |  |  |
| White blood cell count, $\times 10^9/L$ | 4.5 (3.3-5.8) | 5.4 (3.9-7.2) | < .001 |
| Red blood cell count, $\times 10^{12}/L$ | 4.4 (4.1-4.7) | 4.3 (4.0-4.7) | .245 |
| Hemoglobin, g/L | 130.0 (121.0-141.0) | 132.5 (117.0-142.8) | .914 |
| Platelets, $\times 10^9/L$ | 173.0 (140.0-211.0) | 150.5 (117.5-188.8) | .003 |
| Hematocrit, % | 39.5 (36.6-42.7) | 39.4 (36.1-42.7) | .652 |
| Mean corpuscular volume, fL | 90.4 (87.5- 93.4) | 90.5 (87.8-94.3) | .512 |
| Mean corpuscular hemoglobin, pg | 29.9 (28.7-30.9) | 30.2 (29.1-31.4) | .253 |
| Mean corpuscular hemoglobin concentration, g/dL | 330.0 (323.0-336.0) | 330.0 (323.0-336.8) | .709 |
| Red blood cell distribution width standard deviation, fL | 39.1 (36.5-41.0) | 40.1 (14.8-41.9) | .097 |
| Red blood cell distribution width coefficient of variation, % | 12.7 (12.1-14.5) | 13.1 (12.5-38.6) | .004 |
| Platelet distribution width, % | 12.8 (10.7-16.3) | 12.4 (10.7-16.1) | .346 |
| Platelet large cell ratio,% | 24.3 (19.9-30.1) | 25.4 (19.7-31.8) | .192 |
| Mean platelet volume, fL | 9.9 (9.2-10.7) | 10.0 (9.2-10.8) | .146 |
| Thrombocytocrit, % | 0.17 (0.14-0.21) | 0.16 (0.13-0.18) | .009 |
| **White cell differential count** |  |  |  |
| Neutrophil, % | 64.1 (56.6-73.6) | 76.3 (66.1-84.9) | < .001 |
| Lymphocyte, % | 26.2 (19.2-34.8) | 15.8 (8.3-23.2) | < .001 |
| Monocyte, % | 7.7 (5.7-9.8) | 7.0 (4.5-8.8) | .060 |
| Eosinophil, % | 0.2 (0.0-0.6) | 0.0 (0.0-0.3) | .004 |
| Basophil, % | 0.2 (0.1-0.3) | 0.2 (0.1-0.3) | .538 |
| Neutrophil count, $\times 10^9/L$ | 2.9 (1.9-3.9) | 4.1 (2.7-5.6) | < .001 |
| Lymphocyte count, $\times 10^9/L$ | 1.1 (0.8-1.5) | 0.8 (0.5-1.1) | < .001 |
| Monocyte count, $\times 10^9/L$ | 0.3 (0.2-0.4) | 0.4 (0.2-0.5) | .334 |
| Eosinophil count, $\times 10^9/L$ | 0.01 (0.00-0.03) | 0.00 (0.00-0.01) | .040 |
| Basophil count, $\times 10^9/L$ | 0.01 (0.01-0.01) | 0.01 (0.01-0.02) | .424 |
| D-dimer, mg/L | 0.4 (0.2-0.9) | 0.7 (0.4-1.8) | < .001 |
| C-reactive protein, mg/dL | 1.2 (0.4-2.6) | 4.5 (2.4-7.2) | < .001 |
| **Cardiac Enzymes** |  |  |  |
| Aspartate amino transferase, U/L | 21.5 (17.3-30.0) | 32.9 (22.1-42.4) | < .001 |
| Alpha-hydroxybutyric dehydrogenase, U/L | 142.0 (113.0-165.0) | 195.5 (158.0-263.5) | < .001 |
| Lactate dehydrogenase, U/L | 182.0 (141.0-220.0) | 256.0 (216.0-328.4) | < .001 |
| Creatine kinase, U/L | 77.2 (45.0-132.7) | 103.0 (57.8-247.8) | .006 |
| **Liver function** |  |  |  |
| Alanine aminotransferase, U/L | 21.6 (13.3-32.6) | 25.6 (16.0-40.7) | .108 |
| Aspartate transaminase, U/L | 22.1 (17.0-30.4) | 31.1 (21.0-41.6) | < .001 |
| Gamma-glutamyl transpeptidase, U/L | 22.1 (13.3-39.1) | 37.3 (22.0-59.0) | < .001 |
| **Kidney function** |  |  |  |
| Urea, mmol/L | 4.1 (3.2-5.4) | 5.9 (4.5-8.1) | < .001 |
| Creatinine, $\mu\text{mol/L}$ | 63.8 (52.8-76.3) | 76.1 (63.6-90.9) | < .001 |
| Procalcitonin, ng/mL | 0.05 (0.04-0.09) | 0.10 (0.06-0.24) | < .001 |
| B-type natriuretic peptide, pg/mL | 62.2 (23.0-136.2) | 147.3 (35.9-524.0) | < .001 |
| **Electrolyte** |  |  |  |
| Potassium, mmol/L | 4.2 (4.0-4.6) | 4.2 (3.9-4.8) | .836 |
| Sodium, mmol/L | 140.7 (139.0-42.2) | 139.4 (137.5-141.6) | .001 |
| Chloride, mmol/L | 104.0 (102.5-105.5) | 102.6 (101.1-105.3) | .001 |
| Calcium, mmol/L | 2.3 (2.2-2.4) | 2.2 (2.1-2.3) | < .001 |
| Phosphate, mmol/L | 1.1 (1.0-1.3) | 1.0 (0.8-1.3) | .027 |

Data are median (IQR). p values comparing non-severe and severe groups were obtained using the Mann-Whitney U test.

Among the clinical features, age, hypertension, hospital employment, body temperature and the time of onset to admission were selected. Lymphocyte (proportion), neutrophil, (proportion), CRP, lactate dehydrogenas (LDH), creatine kinase (CK), urea and calcium were selected from the laboratory feature set. Only the lesion range score was selected from CT semantic features. When putting these three category features together to select features, age, Lymphocyte (proportion), CRP, LDH, CK, urea and calcium were finally included in the combination model.

Model performance was as follows. The Model 1 based on age, hypertension, and hospital employment showed an AUC of 0.774 (95% CI, 0.711-0.837) on the training dataset and an AUC of 0.839 (95% CI, 0.741-0.937) on the validation dataset. The Model 2 with the clinical features of age, hypertension, hospital employment, body temperature, and the time of onset yield an AUC of 0.789 (95% CI, 0.728-0.849) on the training dataset and an AUC of 0.801 (95% CI, 0.687-0.915) on the validation dataset. The Model 3 based on age and lesion range score on CT, had an AUC of 0.768 (95% CI, 0.700-0.835) on the training dataset and an AUC of 0.873 (95% CI, 0.784-0.962) on the validation dataset. When pooling these three categories of features, the combination model (Model 4) selected 7 features (age, lymphocyte (proportion), CRP, LDH, CK, urea, and calcium), which achieved the highest AUC of 0.866 (95% CI, 0.818-0.914) on the training dataset and an AUC of 0.896 (95% CI, 0.813-0.980) on the validation dataset. The AUC value of Model 4 was significantly higher than Model 1 (p < .001), Model 2 (p < .001), and Model 3 (p = .001) on the training dataset. The median and 25% IQR of prognostic probability on the training is 0.158 and 0.086, the cut-off values of low, medium, and high-risk probabilities were 0.072 (0.158 minus 0.086) and 0.244 (0.158 plus 0.086).

Model 4 based was validated on the six test datasets, which showed AUCs ranging from 0.816 to 0.976 with accuracies ranging from 70.8% to 93.8% (**Table 3**). The ROC, confusion-matrix, and calibration plots are shown in **Figure 3**. The number of severe cases in low, medium and high-risk group on the six test datasets is shown in **Figure 4**. The results of randomizing the outcomes and re-running the analysis yielded AUC of 0.530 for the Model 4.

**Table 3.**
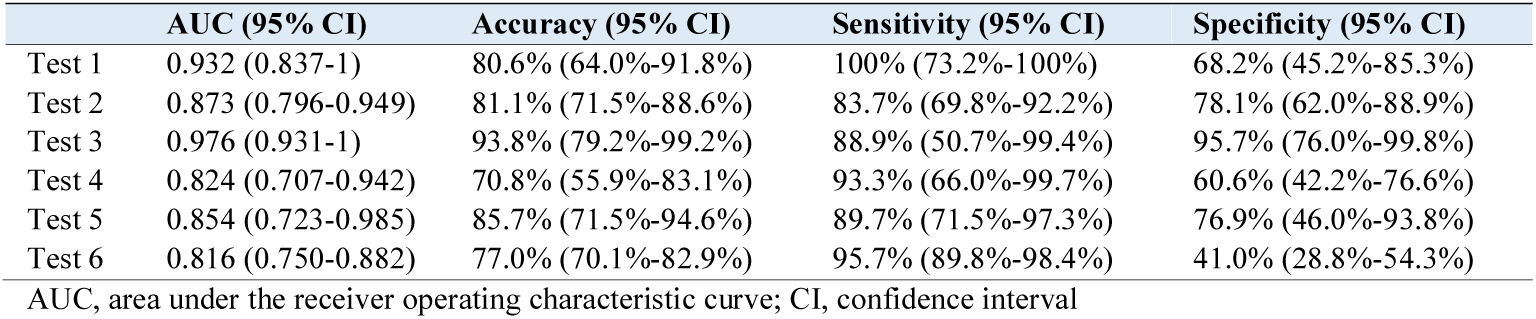
The prognostic performance of the combination model (Model 4) on six test datasets

**Figure 3.**
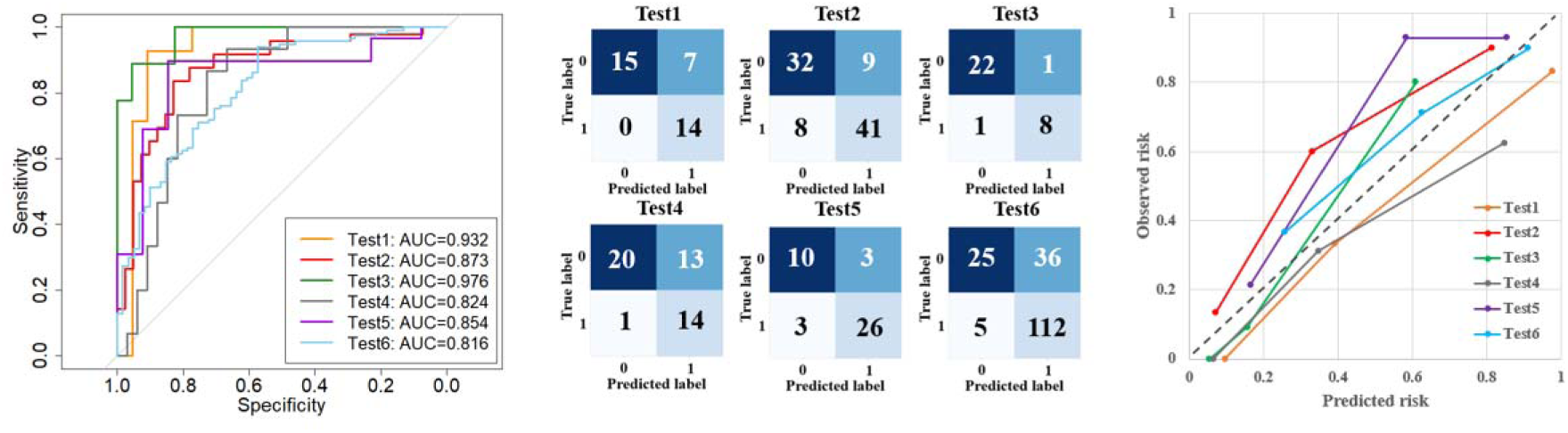
The receiver operator characteristic curve, confusion matrix, and calibration curve for the test datasets

**Figure 4.**
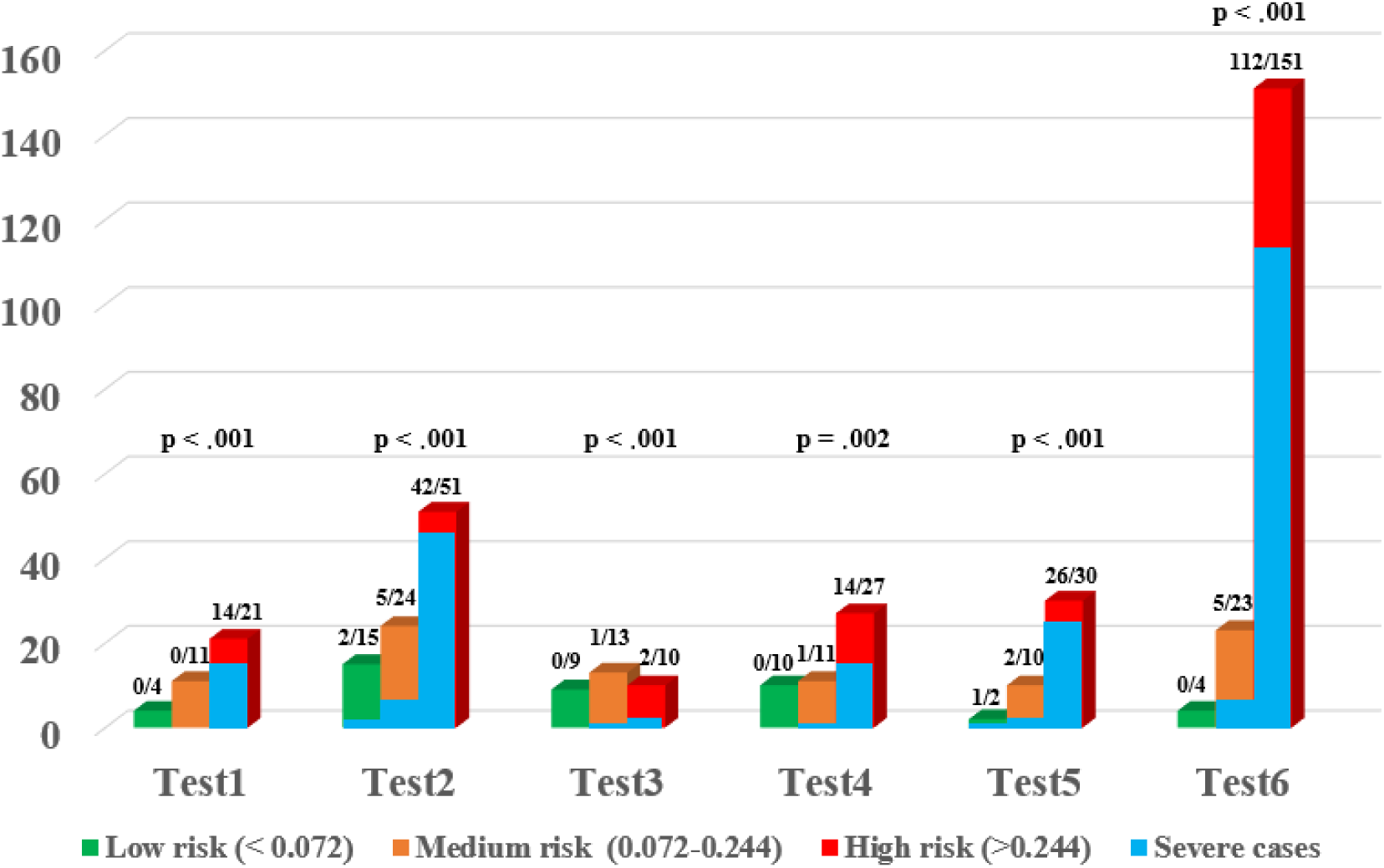
A histogram plot of severe cases in low, medium, and high-risk groups of the test datasets

Based on the selected features from the four models, four different nomograms were established to quantitatively assess the severity risk of illness. These nomograms were available for asymptomatic people, symptomatic patients, radiologists, and clinicians, respectively (**Figure 5**). The developed online-calculators can be found at https://covid19risk.ai/.

**Figure 5.**
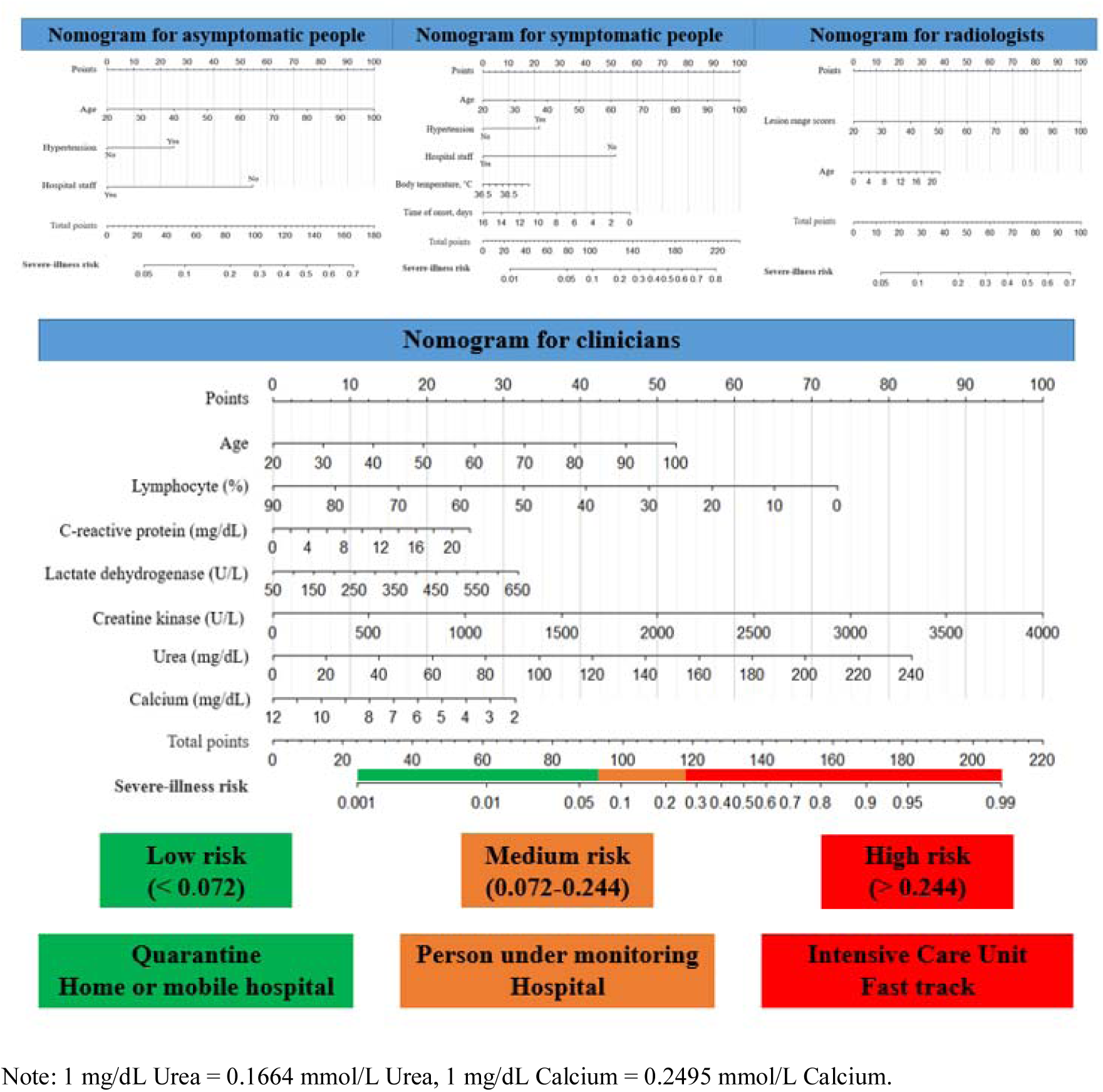
Severe-illness risk nomograms and triage tool for asymptomatic people, symptomatic people, radiologists, and clinicians

## Discussion

This international multicenter study analyzed individually and in combination, clinical, laboratory and radiological characteristics for COVID-19 patients at hospital admission, to retrospectively develop and prospectively validate a prognostic model and tool to assess the severity of the illness and its progression. We found that COVID-19 patients with a severe illness were often of an advanced age, accompanied by multiple comorbidities, presenting with chest tightness, and had abnormal laboratory results and broader lesion range on lung CT on admission. Using simpler linear regression models, which yield similar excellent prognostic performance both in the training and in test datasets. We believe these models could be useful for risk assessment and triage.

Previous studies have reported that age and underlying comorbidities (such as hypertension, diabetes, and cardiovascular diseases) may be risk factors for the COVID-19 patients requiring intensive care unit (ICU).^16,17^ In this study, we found that the elderly COVID-19 patients who were male, non-hospital staff, suffering from hypertension, diabetes, cardiopathy disease, chronic obstructive pulmonary disease, cerebrovascular disease, renal disease, hepatitis B virus infection, surgical history, and chest tightness were more vulnerable to develop a severe illness in the early stages of the disease. Among these features, age, hypertension, hospital staff, body temperature and the time of onset to admission had certain prognostic abilities. Age was the most important feature, which may interact with other features, which was why only age was selected into our combination model (Model 4). Zhou and colleagues.^18^ have confirmed that SARS-CoV-2 uses the same cell entry receptor (angiotensin-converting enzyme II [ACE2]) with SARS-CoV. However, whether COVID-19 patients with hypertension have higher severe illness risk, which is due to treatment with ACE2- increasing drugs is still unknown.^19^ Hospital staff had a lower risk, which may indicate that medical knowledge protects against COVID-19, although the unbalanced nature of this type of data has to be taken into account.

Furthermore, early studies have shown that COVID-19 patients with severe illness had more laboratory abnormalities such as CRP, D-dimer, lymphocyte, neutrophil, and LDH, than those patients with non-severe illness, which were associated with the prognosis.^16,17,20^ In our study, we also found that the severe group had numerous laboratory abnormalities in complete blood cell count, white cell differential count, D-dimer, CRP, liver function, renal function, procalcitonin, B-type natriuretic peptides, and electrolytes. Among these abnormalities, lymphocyte proportion, neutrophil proportion, CRP, LDH, CK, urea, and calcium were significant prognostic factors, which suggest that COVID-19 may cause damage to multiple organ systems when developing into a severe illness. However, current pathological findings of COVID-19 suggest that there is no evidence that SARS-CoV-2 can directly impair the other organs such as liver, kidney and heart.^21^

Current reports have shown that thin-slice chest CT is a powerful tool in clinical diagnosis due to the high sensitivity and the ability to monitor the development of the disease.^22,23^ In addition, a previous study reported that ground-glass opacity and consolidation were the most common CT findings for COVID-19 patients with pneumonia, while being nonspecific.^24^ Clinical observations showed that there were significantly more consolidation lesions in ICU patients on admission, while more ground-glass opacity lesions were observed in non-ICU patients.^25^ In our study, we found that vascular enlargement, air-bronchogram, and lesion range score differ significantly between non-severe and severe groups. Among these features, only the lesion range score had prognostic power, but not enough to be selected for the combination model. This indicates that while these early stage CT semantic features could have diagnostic value, they have limited ability to prognose the onset of severe illness in COVID-19 patients.

The Chinese National Health Committee added some warning indicators for severe or critical cases in the updated diagnosis and treatment plan for COVID-19 patients (version 7),^26^ which includes progressive reduction of peripheral blood lymphocytes, a progressive increase of IL-6, CRP and lactate, and rapid progression of lung CT findings in a short period. In this study, we used age, lymphocyte fraction, CRP, LDH, CK, urea, and calcium scores from clinical, laboratory, and radiological exams recorded at hospital admission to train a model for the prediction of the onset of severe illness. Our model combining these features from multiple sources showed a favorable performance when validated in the six external datasets from China, Italy, and Belgium. In addition, the model is able to stratify COVID-19 patients into low, medium, and high-risk groups for developing severe illness. We propose that this model with its high sensitivity lower specificity could be used for a preliminary screening and triage tool at hospital admission for the potential to develop severe illness. Furthermore, the model could be used for the selection and/or stratification of patients in clinical trials in order to homogenize the patient population. Follow-up laboratory tests are needed to assess the severity risk with a higher accuracy.

As one of the coronaviruses family infecting humans, SARS-CoV-2 has similar etiologic, clinical, radiological and pathological features to those of severe acute respiratory syndrome coronavirus (SARS) and Middle East respiratory syndrome coronavirus (MERS).^20,27,28^ Therefore, we believe that developing a reliable early warning model based on presently clinical, radiological, and pathological data is necessary for current outbreaks and possible future outbreaks of coronaviruses.

### Limitations

Our study has several limitations. First, selection bias is unavoidable due to its retrospective modeling and the limited and unbalanced sample size. Second, patients from different races and ethnicities may have diverse clinical and laboratory results, and the self-medication of patients before admission may affect the clinical and laboratory results. Third, the threshold to go to the hospital can vary from country to country, we are also aware that RNA viruses can mutate rapidly and that could have an impact of the performance of the models. We therefore propose that those models should be continuously updated for example using privacy-preserving distributed learning approaches.^29,30^ Fourth, the CT features used for this study are semantic features from the first CT scan, and quantitative features automatically extracted from CT images using radiomics or deep learning approaches may improve its prognostic performance, and follow-up CT scan may yield more information. Finally, there is also the fundamental weakness of nomograms, which do not give a confidence interval to the final output.

### Conclusion

Elderly COVID-19 patients with hypertension and non-hospital staff seem more vulnerable to develop a severe illness as per defining criteria, which can cause a wide range of laboratory and CT anomalies. Furthermore, our model based on lactate dehydrogenase, C-reactive protein, calcium, age, lymphocyte proportion, urea, and creatine kinase might be a useful preliminary screening and triage tool for risk assessment of patients at hospital admission.

## Data Availability

After the publication of study findings, the data will be available for others to request

## Author Contributors

G. Wu, P. Yang, Y. Xie, X. Wang, and P. Lambin conceived and designed the study. G. Wu and P. Yang contributed to the literature search. P. Yang, X. Rao, J. Li, J. Li, D. Du, S. Zhao, Y. Ding, B. Liu, W. Sun, F. Albarello, A. D’Abramo, V. Schininà, E. Nicastri, J. Wu, M. Occhipinti, G. Barisione, E. Barisione, J. Guiot, A. Frix, M. Moutschen, R. Louis, P. Lovinfosse, and C. Yan contributed to data collection. G. Wu, H. Woodruff, and P. Lambin contributed to data analysis. G. Wu, H. Woodruff, and P. Lambin contributed to data interpretation. G. Wu and C. Yan contributed to the tables and figures. G. Wu, I Halilaj, and P. Lambin contributed to build a website. G. Wu, P. Yang, H. Woodruff, and P. Lambin contributed to writing of the report.

## Supplement

### Test datasets

Test 1: from China Resources Wuhan Iron and Steel Hospital, Wuhan, China (non-severe patients: 22, severe patients: 14). Test 2: from Huangshi Central Hospital, Huangshi, China (non-severe patients: 41, severe patients: 49). Test 3: from Shaoyang Central Hospital, Shaoyang, China (non-severe patients: 14, severe patients: 6), Southern Hospital of Southern Medical University, Guangzhou, China (non-severe patients: 5, severe patients: 1), and Affiliated Zhongshan Hospital Dalian University, Dalian, China (non-severe patients: 4, severe patients: 2). Test 4: from National Institute for Infectious Diseases – IRCCS, Roma, Italy (non-severe patients: 33, severe patients: 15). Test 5: from IRCCS Ospedale Policlinico San Martino, Genoa, Italy (non-severe patients: 13, severe patients: 29). Test 6: from CHU of Liège, Liège, Belgium ((non-severe patients: 61, severe patients: 117)

### CT acquisition and reconstruction parameters

Chest CT scans were performed using one of the CT scanners (uCT 780, United Imaging, China and Brilliance iCT 128, Philips Medical Systems, the Netherlands) with patients in the supine position. The scanning range was from the level of the upper thoracic inlet to the inferior level of the costophrenic angle. For CT acquisition, the tube voltage was 120kVp with automatic tube current modulation, a field of view (FOV) of 350 × 350 mm, and a matrix size of 512 × 512. All images were reconstructed into a slice thickness of 1 mm and an interval of 1 mm.

